# Gender and COVID-19 Vaccine disparities in Cameroon

**DOI:** 10.1101/2022.06.12.22276293

**Authors:** Adidja Amani, Tatiana Mossus, Fabrice Zobel Lekeumo Cheuyem, Chanceline Bilounga, Pamela Mikamb, Jonas Basseguin Atchou, Aude Perine Minyem Ngombi, Armanda Nangmo, Yannick Kamga, Georges Bediang, Joseph Kamgno, Anne-Cécile Zoung-Kanyi Bissek

## Abstract

Six months following the national launch of COVID-19 vaccination in Cameroon, only 1.1% of the target population was fully vaccinated with women representing less than one-third of the vaccinated population regardless of their age, profession or comorbidities. Hence, the aim of this study was to understand the low COVID-19 vaccination rate of women in order to enhance vaccine uptake. A cross-sectional study was conducted between July and October 2021 through an online survey. Also, a retrospective analysis of the Cameroon Ministry of Public Health (MINSANTE) database of the pandemic (COVID-19), for the period of March 2020 to October 2021 was equally carried out. Our sample consisted of 249 responders aged between 18 and 50 years enrolled in the 10 regions of Cameroon, with 142 (57%) who were female. We assessed factors related to having been vaccinated against Covid-19 and predictors to Covid-19 vaccination among non-vaccinated people. Concerning Covid-19 vaccination, 39.2% were not vaccinated. Non-vaccination was statistically associated with being female, being a healthcare worker, fear of adverse effects, and not believing in the vaccine. In the qualitative analysis, women identified themselves as being anti-COVID vaccine for several reasons, including doubts about the quality or safety of the vaccine; the perception that anti-COVID-19 vaccines are presented as being an obligation; including the multitude of vaccines on the market, the believe that there are “more local” effective alternatives to the vaccine. The implementation of the gender approach to COVID vaccination is a condition for the effectiveness and sustainability of actions.

## Introduction

The beginning of the year 2020 was stroked by a worldwide emerging infectious disease called COVID-19. The world has tried to fight against this sickness with several established hygienic and barrier measures to no avail. However, there were still thousands of deaths in most countries of the world. But thanks to the relentless efforts of pharmaceutical companies, at the end of the year 2020 some vaccines against the disease were made available and approved for mass vaccination of populations. Since then, Health authorities have initiated vaccination in their countries even though many are still lagging behind.

Cameroon’s national vaccination campaign was launched on April 12, 2021, with two types of vaccines: Sinopharm and AstraZeneca (1) with an aim to vaccinate at least 15 million people; a number required to achieve in-country herd immunity. Thirty days after the campaign was launched in Cameroon, it was unfortunate to notice women constituted only one third of the vaccinated population regardless of age and health conditions and type of vaccine. Cameroon demographics of 2020 (2) showed that the male to female ratio was 100.06/100 which represents almost half of the total Cameroonian populations so having women not vaccinated might really jeopardize the COVID-19 vaccination rates in Cameroon. The aim of this work was to sort out reasons for this disparity and non-vaccination among Cameroonians and more specifically among women.

## Methodology

### Ethical considerations

The Ethical Review Board of the Faculty of Medicine and Biomedical Sciences of the University of Yaoundé I approved the study. In addition, we received an administrative authorization from the Ministry of public Health of Cameroon. All procedures performed in studies involving human participants were in accordance with the ethical standards of the institutional review board of the committee and with the 1964 Helsinki declaration and its later amendments or comparable ethical standards.

Informed consent was obtained from all individual participants included in the study. The survey started with a consent statement and participants who gave consent to willingly participate in the survey would click the ‘Continue’ button and be directed to complete the self-administered questionnaire. Respondents were free to terminate the survey at any time and no identifying information was captured.

### Study design, period and setting

For the web survey, a cross-sectional observational study was used. It was conducted in Cameroon between July and October 2021. Cameroon is a country in Central Africa with an estimated population in 2020 of 26,545,864 among which 13,268,789 are women (2–4). According to the WHO, there are nearly 1.1 doctors and 7.8 nurses and midwives per 10,000 inhabitants (5). The country is divided into ten regions with heterogeneous socio-demographic characteristics.

A retrospective analysis of the Cameroon Ministry of Public Health (MINSANTE) database of the pandemic (COVID-19), for the period of March 2020 to October 2021 was equally carried out to highlight epidemiological data of the disease, since the beginning of the pandemic in the country.

### Study population

Cameroonian residents, aged 18 years or more, employed or unemployed who understood the content of the form and who agreed to participate in the study completed the questionnaire. Also, all participants whose data were recorder in the COVID-19 database of MINSANTE were included in this study.

### Data collection tools and procedures

The online survey was anonymous, administered in official languages (French and English), and hosted on Google Forms (Alphabet Inc., California, USA). Data was collected using a pre-tested questionnaire (6). This method was chosen because of its logistical advantages and its ease of use, especially in the context of restrictions due to the COVID-19 pandemic. Google Forms is a cloud-based data management tool used to design and develop web questionnaires. This tool is provided by Google Inc10. From the designed form, a web URL address was generated. The link was sent to the different authors of the article who in turn shared it to different participants of the study. In order to reach as many participants as possible, the web link was shared via WhatsApp (Facebook Inc., California, USA), Facebook Inc, and Email (7,8). Data collection complied with the terms and conditions of Google Forms.

Data on COVID-19 pandemic in the country were recorded on daily basis across the national territory from various COVID-19 care centres disseminated throughout the country in a web platform called DHIS 2 (District Health Information Software) used by the Cameroon MINSANTE for national data collection of all programs and easily made available for extraction and analysis at all level of the health pyramid.

### Measures

The form of the web survey consisted of four sections: a briefing note with informed consent, socio-demographic characteristics, determinants and suggestions. The briefing note included a brief introduction on the background, purpose, voluntary characteristic of participation, anonymity and confidentiality statements. The 27 questions of the form were distributed over the other three sections, namely ten elements on socio-demographic characteristics, 14 on the determinants of vaccination against COVID-19 and three elements of suggestions to increase vaccination coverage.

In the database of Cameroon MINSANTE, sociodemographic, clinical, paraclinical and prognostic data of COVID-19 patients were recorded in the DHIS2.

### Data procession and analysis

Once the web questionnaire was completed online, the data was saved to a Google spreadsheet in an analysable format (7). However, the data was extracted into an Excel 2010 (Microsoft Corp., Washington, USA) sheet, where data curation and coding were carried out and imported into SPSS (Statistical Package for Social Science) version 26 software (IBM, New York, USA) to analyse and design different cross tables. The statistical difference between values of cells were assessed using a bilateral test of Fisher exact test. Only statistically significant variables (*p*-value 0.05) in the univariate logistic regression were included in the multivariate logistic regression to establish the strength of an association and control for possible confounding. Depending on whether one was in a univariate or multivariate analysis, crude or adjusted odds-ratios were presented respectively using the Wald test.

Data of the database were extracted from DHIS2 to an Excel 2010 form and then exported to SPSS Version 26 software for further processing and analysis. The threshold of significance was set at 0.05 with confidence intervals (CI) at 95%. A qualitative analysis was done to assess socio-anthropologic roots of vaccinal hesitancy among participants.

## Results

### National epidemiological results on COVID-19 cases and death

From the start of the epidemic in March 2020 until October 2021, 79,861 confirmed cases of COVID-19 were recorded in the national database. The median age was 35 years with an interquartile range of 23 to 40 years. They were mostly adults with the most represented age group 30 to 49 years old with 38.3% followed by that of 50 years and over (Table 1).

**Table 1.** Distribution of confirmed cases of COVID-19 by age group, Cameroon, 2021.

| Age groups (years) | Frequency (%) |
| --- | --- |
| *(Number of positive cases = 79,861)* |  |
| 0-4 | 7923 (9.9) |
| 5-15 | 4381 (5.5) |
| 16-29 | 17418 (21.8) |
| 30-49 | 30596 (38.3) |
| 50+ | 19543 (24.5) |
| Total | 79861 (100) |

On a total of 76,434 confirmed cases in which the sex was known, it appears that the men were more affected than the women with a 50.3% and 45.4% respectively yielding a sex ratio of 1.1. Among affected women with COVID-19, 0.4% were pregnant.

Out of 50,580 cases whose evolution was recorded, 1,232 deaths were notified with a case fatality rate of 2.4%. The risk of death was slightly higher in women than in men with an OR of 1.135 (CI: 1.012-1.274). Regarding age, 71.4% of deaths occurred in people aged 50 and over. The probability of death was 4.6 times higher in the elderly and this difference was statistically significant (p = 0.000). Case confirmation could be made in 482 people who died at the time of notification, accounting for 0.8% (Table 2).

**Table 2.** Distribution of COVID-19 deaths according to age, Cameroon, 2021.

| Variable | Deceased |  |  |  |  |  |
| --- | --- | --- | --- | --- | --- | --- |
|  | *(Number of deceased cases=50 580)* |  |  |  |  |  |
| Age | No | Percentage | Yes | Percentage | Total | Percentage |
| 0-4 | 3386 | 6.9 | 51 | 4.1 | 3437 | 6.8 |
| 5-15 | 2948 | 6.0 | 03 | 0.2 | 2951 | 5.8 |
| 16-29 | 11467 | 23.2 | 49 | 4.0 | 11516 | 22.8 |
| 30-49 | 20026 | 40.6 | 245 | 20.1 | 20275 | 40.1 |
| 50+ | 11521 | 23.4 | 880 | 71.4 | 12401 | 24.5 |
| Total | 49348 | 100 | 1232 | 100 | 50580 | 100 |

### COVID-19 cases and comorbidities

Table 3 indicates that women had more comorbidities than men, that is 6.6% and 5.4% respectively, with an OR of 1.245 (CI: 1.173-1.322). The elderly had more comorbidities than the young people. The majority of people with comorbidities were aged 50 and over (59.8%) and this difference with the young population is statistically significant (p = 0.000).

**Table 3.** Distribution of cases by age and presence of comorbidity, Cameroon 2021.

| Variable | Comorbidity<br>(Number of positive cases=79861) |  |  |  |  |  |
| --- | --- | --- | --- | --- | --- | --- |
|  | Yes | Percentage | No | Percentage | Total | Percentage |
| 0-4 | 112 | 2.5 | 7811 | 10.4 | 7923 | 9.9 |
| 5-15 | 96 | 2.1 | 4285 | 5.7 | 4381 | 5.5 |
| 16-29 | 396 | 8.7 | 17022 | 22.6 | 17418 | 21.8 |
| 30-49 | 1233 | 27.0 | 29362 | 39.0 | 30595 | 38.3 |
| 50+ | 2728 | 59.8 | 16816 | 22.3 | 19541 | 24.5 |
| Total | 4565 | 100 | 75296 | 100 | 79861 | 100 |

Out of the 46,587 forms filled in on health personnel, 3,461 (7.4%) of them were tested positive. Among healthcare workers affected by COVID-19, 25 of them died with a resulting case fatality rate of 0.7% (Table 4).

**Table 4.** Distribution of death among health care workers, 2021.

| Variable |  | Health care worker (n=46 587) |  |  |  |  |
| --- | --- | --- | --- | --- | --- | --- |
| Deceased | No | Percentage | Yes | Percentage | Total | Percentage |
| No | 42614 | 98.8 | 3436 | 99.3 | 46048 | 98.8 |
| Yes | 512 | 1.2 | 25 | 0.7 | 537 | 1.2 |
| Total | 43126 | 100 | 3461 | 100 | 46587 | 100 |

### Web survey results on COVID-19 disparities

#### Demographic characteristics

In total, 249 out of 583 subjects who were approached completed the online survey, yielding a response rate of 42.71%. Among 249 respondents enrolled from the 10 regions of Cameroon, 142 (57%) were female. They were aged between 18 to 50 years old and more with 133 (53.4%) aged between 25 and 34 years old. Majority of respondents were single (45%) followed by married people (41.8%). A total of 130 (52.2%) reside in the Centre Region, and 170 (68.3%) are Health Care Workers (HWC) (Table 5).

**Table 5.** Sociodemographic characteristics of participants (n=249)

| **Variable** | **Number** | **Percentage** |
| --- | --- | --- |
| **Age** |  |  |
| 18-24 | 23 | 9.2 |
| 25-34 | 133 | 53.4 |
| 35-44 | 66 | 26.5 |
| 45-49 | 1 | 0.4 |
| 50+ | 18 | 7.2 |
| **Gender** |  |  |
| Female | 142 | 57.0 |
| Male | 105 | 42.2 |
| Prefer not to say | 2 | 0.8 |
| **Education** |  |  |
| Primary | 2 | 0.8 |
| Secondary | 23 | 9.2 |
| University | 224 | 90.0 |
| **Employment status** |  |  |
| Self employed | 11 | 4.4 |
| Unemployed | 17 | 6.8 |
| Students | 37 | 14.9 |
| Employed | 183 | 73.5 |
| Retired | 1 | 0.4 |
| **Marital status** |  |  |
| Single | 112 | 45.0 |
| Cohabiting | 23 | 9.2 |
| Divorced | 3 | 1.2 |
| Married | 104 | 41.8 |
| Prefer not to say | 7 | 2.8 |
| **Religion** |  |  |
| Catholic | 115 | 46.2 |
| Muslim | 30 | 12.0 |
| Protestant | 78 | 31.3 |
| Other | 26 | 10.4 |
| **Region of residence** |  |  |
| Adamawa | 11 | 4.4 |
| Centre | 130 | 52.2 |
| East | 13 | 5.2 |
| Far-North | 8 | 3.2 |
| Littoral | 26 | 10.4 |
| North-West | 3 | 1.2 |
| North | 27 | 10.8 |
| West | 12 | 4.8 |
| South-West | 10 | 4.0 |
| South | 9 | 3.6 |
| **Having children** |  |  |
| No | 71 | 28.5 |
| Yes | 178 | 71.5 |
| **Health care worker** |  |  |
| No | 79 | 31.7 |
| Yes | 170 | 68.3 |
| **Underlying chronic disease** |  |  |
| No | 228 | 91.6 |
| Yes | 21 | 8.4 |

#### Descriptive epidemiology

This study revealed that nearly 28.91% of participants have experienced sickness due to the novel coronavirus disease. The age group declared to be most affected by COVID-19 was that of 25-34 years old with 38 (15.3%) cases followed by 34-39 years age group and the difference between theses age subgroups were statistically significant (p= 0.043). Among the 249 respondents, females were more affected than men with a proportion of 44 (17.7%) cases. Single and married participants had almost the same proportion of cases estimated at 33 (13.3%) and 31 (12.4%) respectively but with no statistical difference (p= 0.87). People living with children reported more cases of COVID-19 disease (20.9%) than those without children (Table 6).

**Table 6.** Epidemiology of COVID-19 among participants (n=249)

| Variables | Past history of novel coronavirus sickness |  |  |  |  |  | p-value* |
| --- | --- | --- | --- | --- | --- | --- | --- |
|  | Yes | Percentage | No | Percentage | I Don't know | Percentage |  |
| **Age** |  |  |  |  |  |  | **0.043\*\*** |
| 18 – 24 | 8 | 3.2 | 13 | 5.2 | 2 | 0.8 |  |
| 25 – 34 | 38 | 15.3 | 81 | 32.5 | 14 | 5.6 |  |
| 35 – 44 | 25 | 10.0 | 35 | 14.1 | 7 | 2.8 |  |
| 45 – 49 | 0 | 0 | 17 | 6.8 | 1 | 0.4 |  |
| 50+ | 1 | 0.4 | 7 | 2.8 | 0 |  |  |
| **Gender** |  |  |  |  |  |  | 0.42 |
| Female | 44 | 17.7 | 85 | 34.1 | 13 | 5.2 |  |
| Male | 28 | 11.2 | 67 | 26.9 | 10 | 4.1 |  |
| Prefer not to say | 0 |  | 1 | 0.4 | 1 | 0.4 |  |
| **Marital status** |  |  |  |  |  |  | 0.87 |
| Single | 33 | 13.3 | 65 | 26.1 | 14 | 5.6 |  |
| Cohabiting | 7 | 2.8 | 14 | 5.6 | 2 | 0.8 |  |
| Married | 31 | 12.4 | 66 | 26.5 | 7 | 2.8 |  |
| Divorced | 0 |  | 3 | 1.2 | 0 |  |  |
| Prefer not to say | 1 | 0.4 | 5 | 2.0 | 1 | 0.4 |  |
| **Underlying chronic disease** |  |  |  |  |  |  | 0.36 |
| No | 64 | 25.7 | 143 | 57.4 | 21 | 8.4 |  |
| Yes | 8 | 3.2 | 10 | 4.1 | 3 | 1.2 |  |
| **Having Children** |  |  |  |  |  |  | 0.37 |
| No | 20 | 8.1 | 44 | 17.6 | 7 | 2.8 |  |
| Yes | 52 | 20.9 | 109 | 43.8 | 17 | 6.8 |  |

\*bilateral p-value of the Fisher exact test. \*\* <0.05

#### Vaccination status

Out of the 249 study participants, 99 (39.8%) declared having received at least one dose of COVID-19 vaccine. The age group 25-34 years was the most vaccinated group (18.6%) followed by 35-44 years (14.5%). The association between age group and vaccination status was statistically significant (p= 0.001). Majority of participants were not vaccinated (60.2%) and the same age groups above were the most represented. The Female gender was less vaccinated (41.0%) than males. A proportion of 36.1% of participant were vaccinated health care workers and the association between being a HCW and vaccination status was significant (p<0.001). There was a significant association between having children and the vaccination status among the studied population (p= 0.004). People with children and not yet vaccinated being the most represented (39.0%) (Table 7).

**Table 7.** Vaccination status among sociodemographic and clinical groups (n = 249)

| Variables | COVID-19 vaccination |  |  |  | p-value* |
| --- | --- | --- | --- | --- | --- |
|  | No | Percentage | Yes | Percentage |  |
| **Age** |  |  |  |  | **0.001\*\*\*** |
| 18 – 24 | 20 | 8.0 | 3 | 1.2 |  |
| 25 – 34 | 87 | 34.9 | 46 | 18.6 |  |
| 35 – 44 | 31 | 12.4 | 36 | 14.5 |  |
| 45 – 49 | 7 | 2.8 | 11 | 4.4 |  |
| 50+ | 5 | 2.0 | 3 | 1.2 |  |
| **Gender** |  |  |  |  | **<0.001\*\*\*\*** |
| Female | 102 | 41.0 | 40 | 16.1 |  |
| Male | 46 | 18.5 | 59 | 23.1 |  |
| Prefer not to say | 2 | 0.8 | 0 | 0 |  |
| **HCW** |  |  |  |  | **<0.001\*\*\*\*** |
| No | 70 | 28.1 | 9 | 3.7 |  |
| Yes | 80 | 32.1 | 90 | 36.1 |  |
| **Marital status** |  |  |  |  | 0.22 |
| Single | 73 | 29.3 | 39 | 15.7 |  |
| Cohabiting | 16 | 6.4 | 7 | 2.8 |  |
| Married | 55 | 22.1 | 49 | 19.7 |  |
| Divorced | 1 | 0.4 | 2 | 0.8 |  |
| Prefer not to say | 5 | 2.0 | 2 | 0.8 |  |
| **Underlying chronic disease** |  |  |  |  | 0.64 |
| No | 136 | 54.7 | 92 | 36.9 |  |
| Yes | 14 | 5.6 | 7 | 2.8 |  |
| **Having Children** |  |  |  |  | **0.004\*\*\*** |
| No | 53 | 21.3 | 18 | 7.2 |  |
| Yes | 97 | 39.0 | 81 | 32.5 |  |
| **Past history of COVID-19** |  |  |  |  | 0.55 |
| No | 88 | 35.3 | 65 | 26.1 |  |
| Yes | 46 | 18.5 | 26 | 10.4 |  |
| Don't know | 16 | 6.4 | 8 | 3.3 |  |

\*Bilateral p-value of Fisher exact test. \*\* $<0.05$ ; \*\*\* $<0.01$ ; \*\*\*\* $<0.001$

#### Analytic epidemiology

This study noted various factors associated with non-vaccination in the study population. After binary logistic regression analysis, several factors were identified to be significantly associated to non-vaccination. As risk factors: female gender (COR: 3.1; p<0.001), age under 35 years (COR: 2.15; p-value= 0.004), having children (COR 2.64; p=0.002), fear of adverse event (COR: 7.6; p<0.001), no belief in the vaccine (COR: 17.04; p<0.001) and being a HCW. The age between 35-44 years appeared to be protective in univariate analyses probably due to confounders (COR: 0.513; p= 0.019) (Table 8).

**Table 8.**
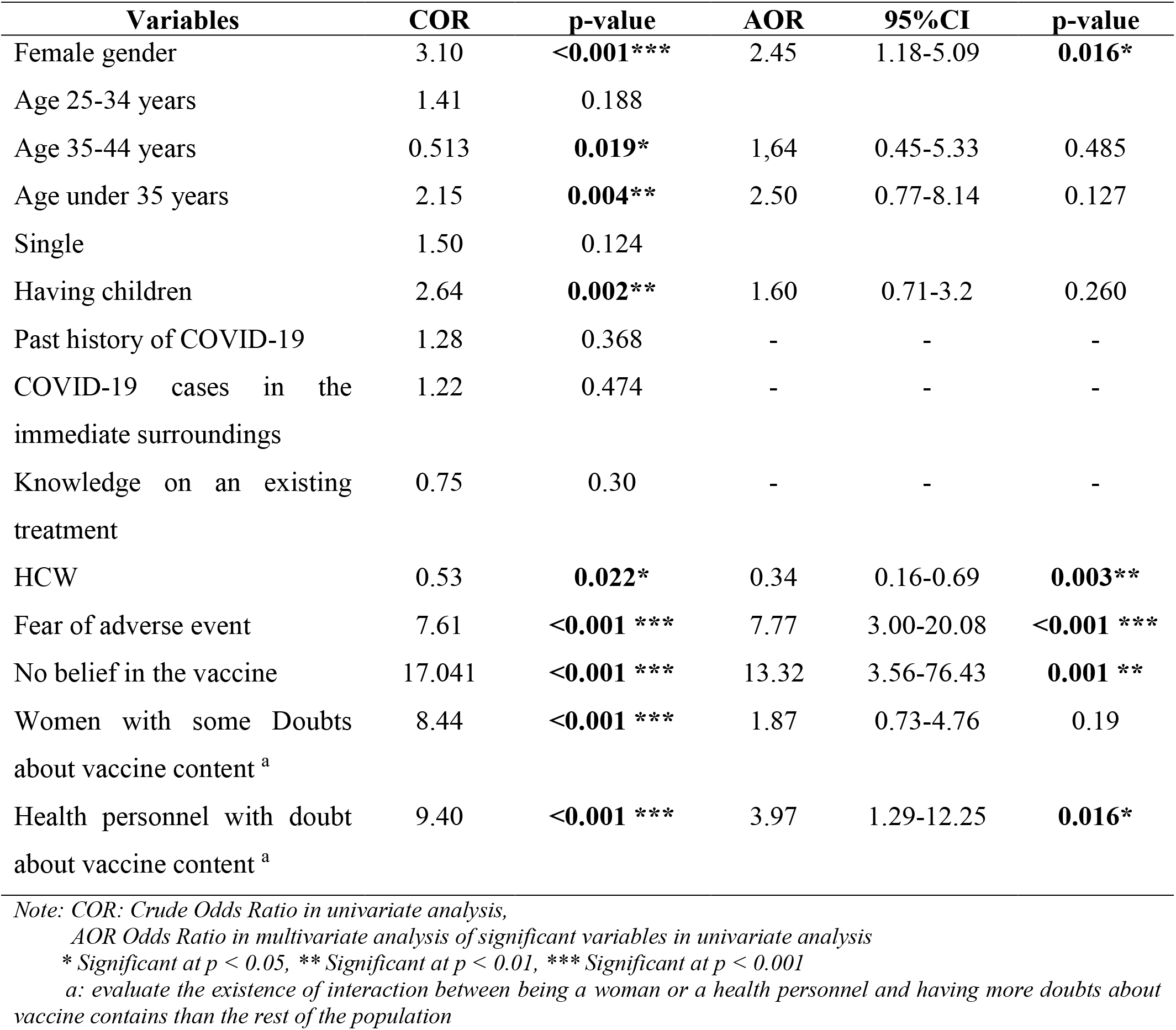
Global synthesis of associated risk factors to COVID-19 non vaccination (n= 249)

| **Variables** | **COR** | **p-value** | **AOR** | **95%CI** | **p-value** |
| --- | --- | --- | --- | --- | --- |
| Female gender | 3.10 | **<0.001\*\*\*** | 2.45 | 1.18-5.09 | **0.016\*** |
| Age 25-34 years | 1.41 | 0.188 |  |  |  |
| Age 35-44 years | 0.513 | **0.019\*** | 1,64 | 0.45-5.33 | 0.485 |
| Age under 35 years | 2.15 | **0.004\*\*** | 2.50 | 0.77-8.14 | 0.127 |
| Single | 1.50 | 0.124 |  |  |  |
| Having children | 2.64 | **0.002\*\*** | 1.60 | 0.71-3.2 | 0.260 |
| Past history of COVID-19 | 1.28 | 0.368 | - | - | - |
| COVID-19 cases in the immediate surroundings | 1.22 | 0.474 | - | - | - |
| Knowledge on an existing treatment | 0.75 | 0.30 | - | - | - |
| HCW | 0.53 | **0.022\*** | 0.34 | 0.16-0.69 | **0.003\*\*** |
| Fear of adverse event | 7.61 | **<0.001 \*\*\*** | 7.77 | 3.00-20.08 | **<0.001 \*\*\*** |
| No belief in the vaccine | 17.041 | **<0.001 \*\*\*** | 13.32 | 3.56-76.43 | **0.001 \*\*** |
| Women with some Doubts about vaccine content <sup>a</sup> | 8.44 | **<0.001 \*\*\*** | 1.87 | 0.73-4.76 | 0.19 |
| Health personnel with doubt about vaccine content <sup>a</sup> | 9.40 | **<0.001 \*\*\*** | 3.97 | 1.29-12.25 | **0.016\*** |

*Note: COR: Crude Odds Ratio in univariate analysis,*
*AOR Odds Ratio in multivariate analysis of significant variables in univariate analysis*
*\* Significant at $p < 0.05$ , \*\* Significant at $p < 0.01$ , \*\*\* Significant at $p < 0.001$*
*a: evaluate the existence of interaction between being a woman or a health personnel and having more doubts about vaccine contains than the rest of the population*

There was an interaction between female gender, being a HCW and having some doubts about the vaccine content. This interaction was shown in bivariate analysis to be statistically significant. For female gender (COR:8.44; p<0.001) and for HCW (COR: 9.40; p=0.001).

Multinomial logistic regression analysis showed that female gender was 2.5 (CI: 1.18-5.09) more at risk of not being vaccinated than men. People experiencing fear of adverse events and with no belief about vaccine content were 7.7 (CI: 3.00-20.08) and 13.32 (CI: 3.56-76.43) times more at risk of not being vaccinated than others. HCW were 2.9 (AOR: 0.53; CI: 0.16-0.69) times less at risk to be unvaccinated. On the other hand, HCW were significantly more susceptible to have doubts about vaccine and those with doubts were 3.91(CI: 1.29-12.25) times more at risk of not being vaccinated than others (Table 8).

#### Female gender specific associated factors

We found that from 142 women enrolled in this study, only 40 (28.2%) were vaccinated against COVID-19. Evaluation of risk factors associated to non-vaccination specifically among women showed the following results. In bivariate logistic regression analysis, women having children (COR: 3.70; p= 0.007), fear of adverse events (COR: 5.41; p= 0.003), doubts about vaccine content (COR:14.61; p<0.001). On the other hand, being a HCW was a favourable factor for the vaccine uptake (COR: 0.098; p<0.001).

After multinomial logistic regression, women having some doubts about vaccine contents were 5.44 times (CI: 1.42-20.86) more at risk of not being vaccinated than those without doubts. Fear of adverse events were also associated to non-vaccination but not statistically significant. Being a female health care worker was 6.6 times (AOR: 0.15; CI: 0.04-0.56) less at risk of not being vaccinated than non HCW women (Table 9).

**Table 9.**
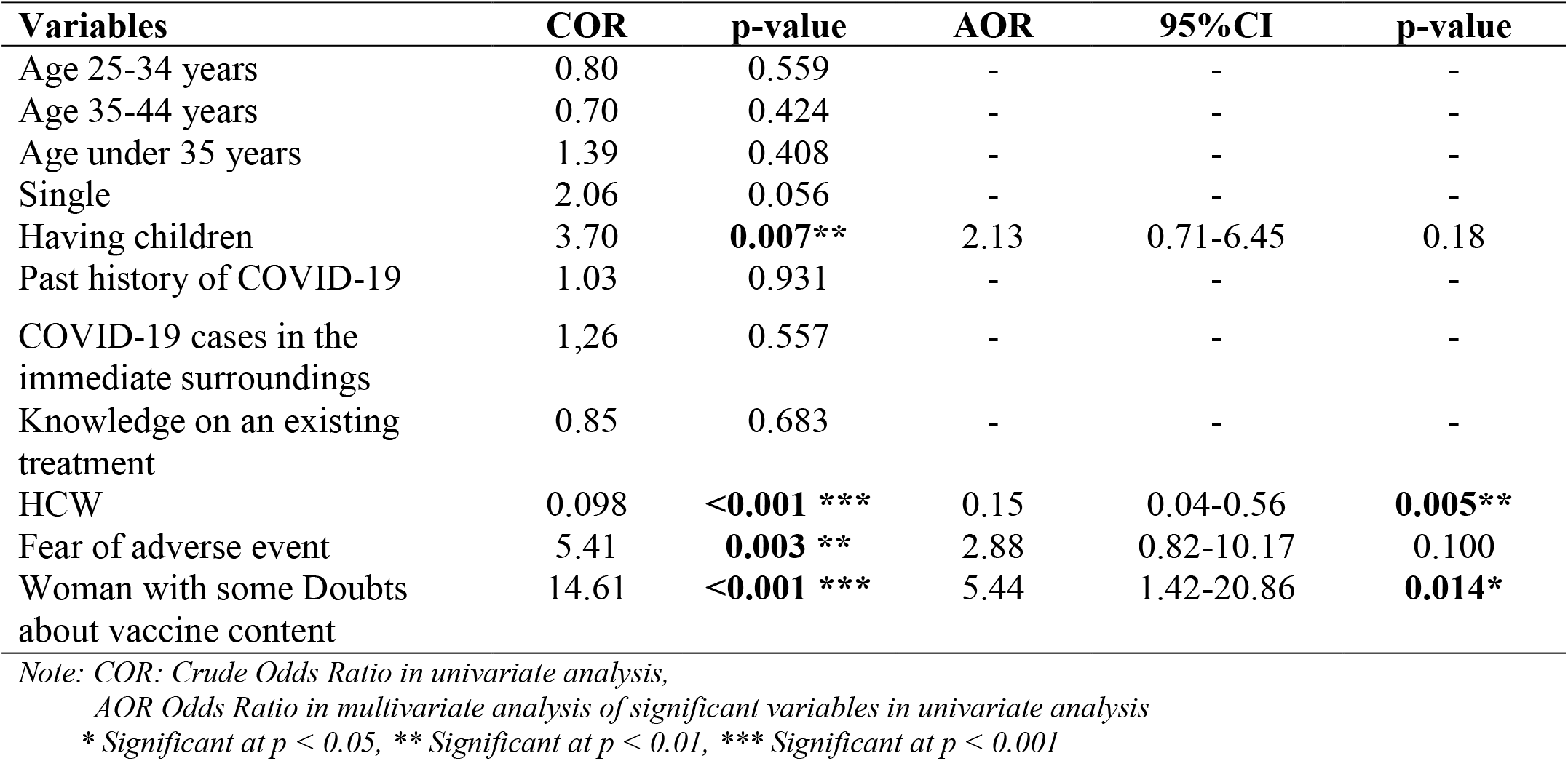
Specific factors associated to non-vaccination among female gender (n= 142)

#### Associated factor to COVID-19 acceptability

Globally, from 150 non vaccinated participants, only 56 (37.33%) had willingness to receive the vaccine. Binary logistic regression revealed that female gender has lower willingness to get the vaccine if they were proposed a COVID-19 vaccine but this result was not statistically significant (COR: 0.65; p= 0.317). Similar findings but statistically significant were noted for those having had COVID-19 cases in the immediate surroundings (COR: 0.18; p= 0.007), no belief in the vaccine (COR: 0.080; p= 0.016), no vaccine preference (COR:0.14; p<0.001) and for women having some doubts about vaccine content (COR: 0.13; p=0.033).

Moreover, results revealed, health professionals (COR: 3.20; p=0.014) and people having Sinopharm vaccine preference (COR: 3.42; p=0.033) have a higher willingness to receive COVID-19 vaccination if they were proposed one (Table 10).

**Table 10.** Predictor of acceptability of later COVID-19 vaccine uptake.

| Variables | COR | P-value | AOR | 95%CI | P-value |
| --- | --- | --- | --- | --- | --- |
| Female gender | 0.65 | 0.317 | - | - | - |
| Age 25-34 years | 0.89 | 0.774 | - | - | - |
| Age 35-44 years | 1.48 | 0.410 | - | - | - |
| Age under 35 years | 0.65 | 0.317 | - | - | - |
| Single | 0.867 | 0.733 | - | - | - |
| Having children | 0.84 | 0.696 | - | - | - |
| Past history of COVID-19 | 1.16 | 0.732 | - | - | - |
| COVID-19 cases in the immediate surroundings | 0.18 | **0.007\*\*** | 0.15 | 0.03-0.56 | 0.007** |
| Knowledge on an existing treatment | 0.82 | 0.667 | - | - | - |
| HCW | 3.20 | **0.014\*** | 1.71 | 0.55-5.31 | 0.350 |
| Fear of adverse event | 0.41 | 0.070 | - | - | - |
| No belief in the vaccine | 0.082 | **0.016\*** | 0.17 | 0.02-1.52 | 0.114 |
| Janssen preference | 3.35 | 0.18 | 1.5 | 0.31-7.33 | 0.611 |
| Sinopharm preference | 3.42 | **0.033\*** | 3.58 | 0.61-21.12 | 0.160 |
| No vaccine preference | 0.14 | **<0.001 \*\*\*** | 0.28 | 0.07-1.12 | 0.072 |
| Woman with some Doubts about vaccine content<sup>a</sup> | 0.13 | **<0.001 \*\*\*** | 0.11 | 0.03-0.411 | **0.001\*\*** |
| Health personnel with doubt about vaccine content<sup>a</sup> | 0.37 | 0.061 | - | - | - |

Note: COR: Crude Odds Ratio in univariate analysis,
AOR Odds Ratio in multivariate analysis of significant variables in univariate analysis
\* Significant at $p < 0.05$ , \*\* Significant at $p < 0.01$ , \*\*\* Significant at $p < 0.001$
a: evaluate the existence of interaction between being a woman or a health personnel and having more doubts about vaccine contains than the rest of the population

Only negative predictor was statistically significant in multinomial logistic regression analysis. It was about people having cases of COVID-19 in the immediate surrounding and women with some doubts about vaccine content who were significantly 6.6 and 9.1 times less susceptible to get vaccinated than others (Table 8).

### Qualitative results analysis

Some informants identified themselves as being anti-COVID vaccines and therefore explained several reasons, including doubts about the quality or safety of the vaccines: “You must first know if the content of the vial is really a vaccine against COVID”; the fact that COVID remains a disease of the “white”; “That they should not force Africans to get vaccinated when the disease kills white people much more”; the fact that the vaccine is perceived or presented as being an obligation: “I am against an imposed vaccine”.

The fact that there are a multitude of vaccines on the market: “The covid-19 pandemic is real but the real problem arises with the variety of vaccines”; “The efficacy difference in percentage of vaccines and different names generally creates doubt in people, especially women”. According to them, women are more insightful than men: “We women have much more wisdom in decision-making. One problem already has several vaccine proposals in such a short time, why? Let the people in charge of those vaccines first agree on one, do some testing and release the results over time, and then we’ll see”. Other informants finally suggest that there are alternatives to the vaccine: “It would be better to use naturopathy, to overcome this pandemic:” Do not take any vaccine, take Ekouk and Mfol, it is effective”.

A few participants had no suggestions to share, either because they had no idea about it or because they felt it was a personal decision: “I have no advice for them. Everyone is free to engage in it or not”. Others, because they themselves, are against the vaccine: “For common vaccines, I can agree. But for COVID-19 I cannot encourage people even less women because, they are responsible for lives and their security”.

The COVID pro-vaccines thought that the vaccination of women should be obvious because: “For me, women are more vulnerable because of their fragility”; “She must protect herself and those around her since women remain at the center of many activities”. One respondent, however, underlined the incongruity of the question because according to him: “There is no difference for me, the same actions must be carried out in both sexes”.

While some informants refrained from contributing to the topic, others were able to offer suggestions for raising the vaccination rate among women. Respondents believe that the solution could come from vaccines, for example by ensuring their reliability, by involving African companies in the manufacture of the COVID-19 vaccine or by manufacturing vaccines for young people and pregnant women. But we could also: “Distribute the same vaccine throughout the country. Through:” Introduction of routine vaccine”. Which means, “We can take advantage of pregnancy to let them be vaccinated”. Without forgetting to “Bring the vaccine closer to the population, that is to say more vaccination centers even in the neighbourhoods. “Because:” The vaccination team does not cover a tenth of health areas”.

Support vaccination, that is: “Provide financial motivation for vaccination. “Or also think about:” Make compensation commitments in case the side effects exist”.

To improve women’s adherence to the COVID-19 vaccination, some informants believed that it was necessary to: “First gain men’s confidence in the admissibility of globally credible and acceptable vaccines”. The majority see in: “Awareness. “or” Communication.” as the appropriate solutions to implement. Upstream: “We must first identify the reasons for their refusal and after that it will be easier to carry out campaigns to raise their awareness. “. In order for women to be more receptive, it would be necessary to: “Engage female leaders at all social levels. “. So: “We need to develop better communication on COVID. “. This can be presented in several ways: “Proximity awareness. “; “Targeted awareness. “; “Door to door awareness. “; “Community sensitization “; “Educational meeting. “; “Put the posters everywhere. “.

More specifically, it is: “th need to make them understand that the vaccine is for both sexes and has no negative effects on women.”; “That, famous women who received the vaccine communicate strongly about it. » and « They need role models. “; “Make videos with more actresses that show the benefits of the vaccine and develop messages that contradict the fake news about vaccination. “;” Raise awareness, testify, remind people of the benefits of vaccination, give the right information on the risks on fertility, procreation, breastfeeding, etc. »; “I have published my photos of the vaccine action in associations and meeting, I do with my neighbourhood comrades. I made them aware of vaccination benefits. “;” Get closer to women’s associations, community meetings for women, put women at the front of the business. “; “Good communication and essential messages with community leaders, religious, influencers and extend to all women layers.”;” Focused sensitization and health education. Encouragement and no attempt at all at coercion or obligation. Government has to increase opportunities for sensitization of women and general public. But also, we have to be honest and produce very credible and convincing reports on how we have used COVID-19 funds, because this issue of mismanagement of COVID funds has come to embolden earlier suspicion that there is no COVID and that government is using it to get money from Europe and America and from monetary organizations. “; “Raise awareness of the importance of the vaccine and reduce beliefs that the vaccine is harmful or lethal. “;” Organize sensitization sessions in health facilities, in places of gathering (places of worship, markets, etc.), explain to men the need to have their girlfriends, wives, sisters, mothers, etc. vaccinated. “; “deny fake news from the social media. “;” Strengthen pre and post vaccination counselling “.

“We recognize the good tree by the sweetness of its fruits, if we realize that the vaccine really protects with contamination drop in vaccinated countries like Israel, sensitization will be easy and convincing. “. Statistics from good student countries such as Israel might just as well be convincing. However, the most drastic advocate: “Make vaccination compulsory. “. On the second question of how to use social media to improve vaccine confidence and vaccine uptake, a few informants had no knowledge to share. Other respondents shared their doubts about the feasibility of this project: “Very difficult because we find too many rumours in social media. “.

Some people thought that social networks were not suitable for impacting the population: “The media and Radio Stations are great sources of manipulation. The population has already understood it, that is why you will always see thousands of likes but few acts. “But according to some people,” Social networks should rather help to sensitize people on the respect of barrier measures. “. To improve confidence in vaccines and vaccine uptake, it is important to: “Do not avoid the mistakes of scientists. One dose, two doses, three doses. Vaccinated people who still get contaminated by the disease etc. “.

Social networks are important channels of communication and networking between communities, for informants, their use in improving confidence in vaccines and vaccines uptake can be defined as: “Communicate regularly on: the role of vaccines, the advantages of prevention over treatment, the quality of the vaccine, the possible side effects and their management in Cameroon, indicate the vaccination points, schedules etc. “; “Post the comparative reports of a country affected by covid19 before and after taking the vaccine. “; “By posting videos of people who have received the vaccines, videos that explain the types of vaccine “; “Aggressively invest social networks with messages, videos and images on the benefits of vaccination. “; “Encourage more social groups to create their accounts such as on Facebook, WhatsApp, Telegram and let the leaders of such groups share their pics on how they already took the jab. “; “Make videos from theatres to illustrate the benefits of COVID-19 vaccination and the harms of not doing so. “; “By live sessions during the vaccination sessions in the sites and share them. “; “Invite vaccinated and infected people to speak at a round table or a health program. “; “By using our status to share positive images. “; “Take pictures showing people who get vaccinated and who give good testimonials. “; “By sending messages to our contacts informing them about the importance of the vaccine, by advertising through leaflets on the benefits of the vaccine. “; “The government itself should have their own specific social media platform, whose duty will be to scan social media for all the COVID vaccine negative propaganda, and elaborate simple, clear, objective and convincing messages, reports, short videos or audios, brief social media slots to disperse the propaganda. To make info available to everybody. Social media is the most followed media today and we must be serious to get on with it”.

It emerges from the respondents that all communication through social media should be done transparently and by professionals, people who have been vaccinated, or opinion leaders, influencers trained for this purpose.

The respondents wish to point out that social networks, although important for information dissemination, professionals should take a closer look at the said information circulating: “A lot of rumours and fake-news are disseminated through social networks”; They offer several solutions to this, including: “Set up at all levels, monitoring / listening committees for social networks.”; “Strengthen the information system and rumour management. “; “To counter information, that is to say deny what has been said with the help of experts in the field. “; “prevent doctors and other health personnel who are against the vaccine to give their opinions on TV channels to discourage others. “; “The state needs to control all the sterile information that can create social psychosis, if possible, completely suppress social media. “; “why not even punish those who spread fake news of the vaccines. “.

## Discussion

### Epidemiology of COVID-19

As of October 2021, Cameroon registered 79861 cases with women being more represented, this could be explained by the high demand of health care among female gender when they feel ill. In contrast, during the initial outbreak, in an epidemiological study of 425 coronavirus cases in Wuhan, 56% were male, and the median infected individual age was 59 years which is more than the one of this study which found a median age of 35 years (9,10). The age group 30-49 years was the most represented among COVID-19 cases. Cameroon being an African country with a young working population, this could explain the fact that they are the most numerous to be contaminated by COVID19. A study carried out in February 2020 found that the 30-70 years old age group represent 86.6% of COVID-19 cases in the world (10). A case fatality rate (CFR) of 2.4% was computed in this study. This result is similar to 2.3% found in a study in February 2020 (10) inferior to the world global CFR of 6.7% found in Africa review in April 2020 (11). Moreover, in Africa, countries such as Egypt, Algeria, Burkina Faso, and Morocco had CFRs above 5% (12). Majority of COVID-19 death among cases were concerning the 50 years old and more group; this could be explained by the fact that this age group is more susceptible to have underlying comorbidities that are risk factor of death due to COVID-19. In addition, our study found that majority of COVID-19 patients with comorbidity were people aged 50 years old and over. We recorded 0.7% of death among health care workers which closes to 0.5% result found in a metanalysis in 2020 (13).

### COVID-19 vaccination disparities

Vaccine hesitancy is an old phenomenon that represents a serious threat to global health, as shown by the resurgence of some infectious diseases (e.g., outbreaks of measles and pertussis)(14,15). The huge leaps in developing efficacious and safe COVID-19 vaccines within a short period were unprecedented (16,17). Nevertheless, COVID-19 vaccine hesitancy can be the limiting step in the global efforts to control the current pandemic with its negative health and socio-economic effects (18,19).

In this study, the aim was to highlight reasons for disparity and non-vaccination among Cameroonians and specifically among the female gender. We found that nearly 39.8% of participants had received at least one dose of COVID-19 vaccine which is much higher than national coverage which is estimated around 4% (20). Some sociodemographic factors, such as sex, age and having children were associated with vaccine uptake, in addition to some psychological factors, especially the perception of adverse events and vaccine contents of COVID-19 vaccines(21). Our findings represent one of the first estimates of uptake and acceptance of a COVID-19 vaccine in Cameroon that can be used to plan COVID-19 vaccine uptake among the general public and especially women target.

Vaccinations are widely recognized as one of the most effective preventive measures in public health(22). Vaccine hesitancy varies across time, place, and type of vaccine, and is influenced by a variety of factors(23). Therefore, it is necessary to assess vaccine acceptance of the COVID-19 vaccine and factors that influence it in each region, in order to plan educational activities to increase vaccine acceptance. Previous studies conducted outside of Cameroon reported that various factors, such as sociodemographic factors, attitude and beliefs regarding COVID-19 infection and vaccination, and political views, influence decision-making of vaccine acceptance(24–28). Our results showed, the majority of participant aged between 25 to 34 years were the most represented, this could be due to the method of enrolment into the study who used social network to share questionnaire and this age group is particularly connected on this media of communication according to Facebook Data(29).

Vaccine acceptance in Cameroon was lower among several sociodemographic groups, such as female gender, adult aged 25-34 years, those having had COVID-19 cases in the immediate surroundings, with no belief in the vaccine, no vaccine preference and for women having some doubts about vaccine content which coincided with many previous studies(21,24,26–28,30). In order to increase COVID-19 vaccine coverage in Cameroon, it may be important to ensure vaccination among these populations with low vaccine acceptance. Therefore, interventions targeted at modifying such health beliefs about COVID-19 may lead to improved vaccination rates.

People having a contact with COVID-19 case has lower willingness to receive the vaccine, different result were found in a USA study where this group of individual had at contrary higher willingness to get the vaccine(31); this suggests that perception if this disease by population would be quite different from one region to another due to sociocultural background. Moreover, other studies found similar results concerning women who had lower willingness to get vaccinated(21,31,32). We found a proportion of 37.73% of participant having willingness to get vaccinated which is lower than finding of studies done in Japan(21) and USA(33). A global survey of 19 countries showed that 71.5% of the participants responded that they would receive a vaccine if it was proven to be safe and effective(30).

A total number of 94(62.67%) participants had a low level of willingness to protect others by getting oneself vaccinated. This may suggest that a certain number of people wants to benefit from the indirect protection provided by the vaccination of other people, without getting vaccinated themselves(21). Therefore, trying to change people’s perceptions of the effectiveness of the COVID-19 vaccine, and increasing awareness to willingly protect others by getting oneself vaccinated, may be important in promoting vaccine acceptance.

### Psychological barriers mentioned in a global and non-specific way

Vaccination hesitation can be attributed to a set of psychosocial factors observable outside the religious context and the current state of health of individuals. A proportion of 60.2 % unvaccinated and 50.2 % without a vaccination plan.

#### Lack of information

misunderstanding and not knowing what to think about the need to be vaccinated. Contradictory opinions and an information deficit can generate some perplexity: why getting vaccinated if, in one way or another, you can catch the virus and transmit it? Why vaccinate young people if they are less vulnerable to the virus? Not finding a satisfactory answer to these questions paralyzes thinking and decreases mobilization.

#### Anxiety and fears related to stings

apprehension of needle or pain is sometimes so anxiety-provoking that it can lead to avoiding any situation directly or indirectly involving the vaccination. To the point where sometimes the mere sight of vaccination pictures can be a source of anxiety. In front of an anxiety-provoking situation, everyone reacts differently. Some will be in action and find solutions, others will confide in relatives, or be more emotionally overwhelmed. Others will react in denial. Denial is an automatic, unconscious reflex that acts as a band-aid to control anxiety. In the pandemic context, this can be expressed as denial of the severity of the disease, denial of one’s own vulnerability to contract the virus, or even denial of the existence of the virus.

#### Feeling of rejection and exclusion

As a social being, we are extremely sensitive to rejection. Some individuals have had a life course in which experiences of rejection have been more present and painful than others. They feel more excluded from society, do not recognize themselves in the official discourse and the proposed standards. According to them, the announced health measures are perceived as “controlling”. When, like them, we feel neither represented nor listened to by the authorities, when we are parodied or criticized by other groups in society, we reactivate the wounds of a past already marked by rejection and we replay them in a current sufferer. The person will also feel more excluded and will be less open to following recommendations. She is also likely to feel better understood by alternative and refractory voices that make her dare to hear that she is finally heard.

#### Dependence and conflict avoidance

Some people (women) are more dependent on the opinions of those close to them (their spouses). Relationship dynamics ensure that the person can doubt himself and trust the other to make day-to-day decisions, while seeking to minimize conflicts with him. This seems to be the case, in particular, for women in the African and Cameroonian socio-cultural context in particular. In these cases, the position and choice of the person will be influenced by the fact that a peer does not consider vaccination to be important.

#### Confidence crisis

Aforementioned factors may crystallize into greater mistrust of government sources, health authorities and the pharmaceutical industry. And also, towards a crisis of confidence and mistrust of what is on offer. Conspiracy theory and the rejection of authority come to shape thought and identity. Some will need more explanation, others to be accompanied when receiving the vaccine, or to have a space to feel listened to and accepted in their sense of irritation. Finally, to avoid feeling “controlled”, people will prefer to follow alternative recommendations such as having regular screening tests - rather than receiving the vaccine. In order to offer relevant solutions and move forward collectively in this pandemic crisis, let’s better understand everyone’s reactions in order to better guide the authorities in the transmission of information, as well as in the choice and presentation of measures. For a measure to be respected, one must know the underlying reasons for its rejection.

### Limitations

There are several limitations in the present study. Firstly, subjects were recruited and surveyed online instead of a face-to-face interview which may lead to a potential risk of selection bias. Hence, people without internet access or who are unwilling to participate in online surveys might still not be reached. Also, during survey implementation, the absence of human interaction can encourage hesitation and make it difficult to differentiate junk mail from actual research. Secondly, psychological patterns linked to vaccine hesitation were general and nonspecific thus, the need for a more specific and in-depth study on the subject.

## Conclusion

This study offers insights into reasons of women falling behind in the race to vaccinate against COVID-19. Concerns about vaccine safety is a major contributing factor. Vaccine equity and gender have a role to play in national health responses to COVID-19. The implementation of the gender approach to COVID vaccination is a condition for the effectiveness and sustainability of actions.

## Data Availability

All data produced in the present study are available upon reasonable request to the authors

## Acknowledgements

This study received no specific funding or grant from any agency in the public, commercial, or not-for-profit sectors.

## REFERENCES

1. Amani A, Dove D, Bita A. The First 30 Days of COVID 19 Vaccination in Cameroon: Achievements, Challenges and Lessons Learned. 2021 Jun 3 [cited 2021 Nov 10]; Available from: https://www.preprints.org/manuscript/202106.0094/v1

2. Population, total - Cameroon | Data [Internet]. [cited 2021 Oct 20]. Available from: https://donnees.banquemondiale.org/indicateur/SP.POP.TOTL?end=2020&locations=CM&start=1960&view=chart

3. Cameroon Population (2021) - Worldometer [Internet]. [cited 2021 Oct 20]. Available from: https://www.worldometers.info/world-population/cameroon-population/

4. Population, femmes - Cameroon | Data [Internet]. [cited 2021 Oct 20]. Available from: https://donnees.banquemondiale.org/indicator/SP.POP.TOTL.FE.IN?end=2020&locations=CM&start=1960&view=chart

5. OMS | Cameroun [Internet]. WHO. World Health Organization; [cited 2021 Oct 20]. Available from: https://www.who.int/workforcealliance/countries/cmr/fr/

6. Kanmounye US, Mbonda AN, Djiofack D, Daya L, Pokam OF, Ghomsi NC. Exploring the knowledge and attitudes of Cameroonian medical students towards global surgery: A web-based survey. Wright JG, editor. PLoS ONE. 2020 Apr 30;15(4):e0232320.

7. Vasantha Raju n, N.S. H. Online survey tools: A case study of Google Forms. In 2016.

8. Ngwewondo A, Nkengazong L, Ambe LA, Ebogo JT, Mba FM, Goni HO, et al. Knowledge, attitudes, practices of/towards COVID 19 preventive measures and symptoms: A cross-sectional study during the exponential rise of the outbreak in Cameroon. PLoS Negl Trop Dis. 2020 Sep 4;14(9):e0008700.

9. Early Transmission Dynamics in Wuhan, China, of Novel Coronavirus–Infected Pneumonia | NEJM [Internet]. [cited 2022 Jan 9]. Available from: https://www.nejm.org/doi/full/10.1056/NEJMOa2001316

10. Rauf A, Abu-Izneid T, Olatunde A, Ahmed Khalil A, Alhumaydhi FA, Tufail T, et al. COVID-19 Pandemic: Epidemiology, Etiology, Conventional and Non-Conventional Therapies. International Journal of Environmental Research and Public Health. 2020 Jan;17(21):8155.

11. Lone SA, Ahmad A. COVID-19 pandemic – an African perspective. Emerging Microbes & Infections. 2020 Jan 1;9(1):1300–8.

12. Fadaka AO, Sibuyi NRS, Adewale OB, Bakare OO, Akanbi MO, Klein A, et al. Understanding the epidemiology, pathophysiology, diagnosis and management of SARS-CoV-2: Journal of International Medical Research [Internet]. 2020 Aug 26 [cited 2022 Jan 8]; Available from: https://journals.sagepub.com/doi/10.1177/0300060520949077?url_ver=Z39.88-2003&rfr_id=ori%3Arid%3Acrossref.org&rfr_dat=cr_pub++0pubmed

13. Gómez-Ochoa SA, Franco OH, Rojas LZ, Raguindin PF, Roa-Díaz ZM, Wyssmann BM, et al. COVID-19 in Health-Care Workers: A Living Systematic Review and Meta-Analysis of Prevalence, Risk Factors, Clinical Characteristics, and Outcomes. American Journal of Epidemiology. 2021 Jan 4;190(1):161–75.

14. Phadke VK, Bednarczyk RA, Salmon DA, Omer SB. Association Between Vaccine Refusal and Vaccine-Preventable Diseases in the United States: A Review of Measles and Pertussis. JAMA. 2016 Mar 15;315(11):1149–58.

15. Benecke O, DeYoung SE. Anti-Vaccine Decision-Making and Measles Resurgence in the United States. Glob Pediatr Health. 2019;6:2333794X19862949.

16. Graham BS. Rapid COVID-19 vaccine development. Science. 2020 May 29;368(6494):945–6.

17. Sharma O, Sultan AA, Ding H, Triggle CR. A Review of the Progress and Challenges of Developing a Vaccine for COVID-19. Front Immunol. 2020;11:585354.

18. Harrison EA, Wu JW. Vaccine confidence in the time of COVID-19. Eur J Epidemiol. 2020 Apr;35(4):325–30.

19. Pogue K, Jensen JL, Stancil CK, Ferguson DG, Hughes SJ, Mello EJ, et al. Influences on Attitudes Regarding Potential COVID-19 Vaccination in the United States. Vaccines (Basel). 2020 Oct 3;8(4):E582.

20. Cameroon: WHO Coronavirus Disease (COVID-19) Dashboard With Vaccination Data [Internet]. [cited 2021 Oct 18]. Available from: https://covid19.who.int

21. Machida M, Nakamura I, Kojima T, Saito R, Nakaya T, Hanibuchi T, et al. Acceptance of a COVID-19 Vaccine in Japan during the COVID-19 Pandemic. Vaccines (Basel). 2021 Mar 3;9(3):210.

22. Andre FE, Booy R, Bock HL, Clemens J, Datta SK, John TJ, et al. Vaccination greatly reduces disease, disability, death and inequity worldwide. Bull World Health Organ. 2008 Feb;86(2):140–6.

23. MacDonald NE. Vaccine hesitancy: Definition, scope and determinants. Vaccine. 2015 Aug 14;33(34):4161–4.

24. Reiter PL, Pennell ML, Katz ML. Acceptability of a COVID-19 vaccine among adults in the United States: How many people would get vaccinated? Vaccine. 2020 Sep 29;38(42):6500–7.

25. Wang J, Jing R, Lai X, Zhang H, Lyu Y, Knoll MD, et al. Acceptance of COVID-19 Vaccination during the COVID-19 Pandemic in China. Vaccines (Basel). 2020 Aug 27;8(3):E482.

26. Leng A, Maitland E, Wang S, Nicholas S, Liu R, Wang J. Individual preferences for COVID-19 vaccination in China. Vaccine. 2021 Jan 8;39(2):247–54.

27. Murphy J, Vallières F, Bentall RP, Shevlin M, McBride O, Hartman TK, et al. Psychological characteristics associated with COVID-19 vaccine hesitancy and resistance in Ireland and the United Kingdom. Nat Commun. 2021 Jan 4;12(1):29.

28. Nguyen KH, Srivastav A, Razzaghi H, Williams W, Lindley MC, Jorgensen C, et al. COVID-19 Vaccination Intent, Perceptions, and Reasons for Not Vaccinating Among Groups Prioritized for Early Vaccination - United States, September and December 2020. MMWR Morb Mortal Wkly Rep. 2021 Feb 12;70(6):217–22.

29. Global social media statistics research summary 2022 [Internet]. Smart Insights. 2021 [cited 2021 Nov 18]. Available from: https://www.smartinsights.com/social-media-marketing/social-media-strategy/new-global-social-media-research/

30. Lazarus JV, Ratzan SC, Palayew A, Gostin LO, Larson HJ, Rabin K, et al. A global survey of potential acceptance of a COVID-19 vaccine. Nat Med. 2021 Feb;27(2):225–8.

31. Kreps S, Prasad S, Brownstein JS, Hswen Y, Garibaldi BT, Zhang B, et al. Factors Associated With US Adults’ Likelihood of Accepting COVID-19 Vaccination. JAMA Netw Open. 2020 Oct 1;3(10):e2025594.

32. T A, Ea A, J O-K, M K, A A, Jh A. Examining Vaccine Hesitancy in Sub-Saharan Africa: A Survey of the Knowledge and Attitudes among Adults to Receive COVID-19 Vaccines in Ghana. Vaccines [Internet]. 2021 Jul 22 [cited 2021 Nov 15];9(8). Available from: https://pubmed.ncbi.nlm.nih.gov/34451939/

33. Szilagyi PG, Thomas K, Shah MD, Vizueta N, Cui Y, Vangala S, et al. National Trends in the US Public’s Likelihood of Getting a COVID-19 Vaccine-April 1 to December 8, 2020. JAMA. 2020 Dec 29;

